# How to evaluate the success of the COVID-19 measures implemented by the Norwegian government by analyzing changes in doubling time

**DOI:** 10.1101/2020.03.29.20045187

**Authors:** Biljana Stangeland

## Abstract

Doubling Time (DT) is typically calculated for growth curves that show exponential growth, such as the cumulative number of COVID-19 cases day by day. DT represents the time it takes before the number of COVID-19 cases, in a certain country or area, doubles.

Throughout the ongoing COVID-19 outbreak, DT values are continually changing. These changes are influenced by the measures that are recommended by the health authorities and implemented by governments.

After the government-imposed shutdowns of Nordic Countries that were announced around the 12^th^ of March 2020, we followed the development of the DT in the region. Governments put in place measures never before experienced during peace time; working from home, closed schools and kindergartens, travel bans and social distancing. We conducted analyses to evaluate the effectiveness of these measures. Does it work? The initial set of results following the shutdown are encouraging, demonstrating a trend towards slower growth; however, this could be reversed if the measures that are in place now are abandoned too early. Premature optimism can be very costly. In this report we describe a method for monitoring the epidemic in real time and evaluating the effectiveness of the implemented measures.

## Introduction

Strict measures implemented in many countries around the world, in order to prevent the spread of COVID-19, have put strains on the economy and immobilized the normal functioning of society. Never before in peace time has the modern world seen widespread travel bans, closed borders, home working, and shutdowns of schools and kindergartens. But do these measures successfully slow down and reduce the spread of COVID-19? Is it possible to follow infection trends in real-time to help government officials make well-informed decisions in order to contain the pandemic? Here we describe a method that allows close monitoring of trends that otherwise may not be apparent based on data from standard statistics of cases. By measuring the increase in doubling time, an indicator of how quickly the number of cases is multiplying, we can gain important insights from the COVID-19 epidemiologic data.

The example of Nordic countries can be used to illustrate how to monitor the trends that can correctly predict outcomes in the near future. The COVID-19 outbreak which started in China in December 2019^1^ spread rapidly to the rest of the world, particularly throughout Western Europe and consequently reaching Nordic countries, by the end of February 2020^2^. Nordic and Scandinavian countries, Finland, Norway, Denmark, Sweden and Iceland, are typically associated with high standards of living, high investment in the public health sector and low population density.

In Norway and Denmark, the first cases of COVID-19 were confirmed on the 26^th^ of February, with Iceland following a few days later^2^. Although Finland and Sweden reported one case each a few weeks earlier, new cases began to increase parallel to Norway and Denmark from the 26^th^ of February. Unlike some central European countries, such as France, Germany and the UK, which reported moderate spreading of COVID-19 during February, the number of cases in Scandinavian countries had grown rapidly from the very start (Figure 1A). By the beginning of March, Iceland and Norway were topping the European statistics with the highest infection rates (number of confirmed cases per number of inhabitants)^3^. Two weeks later, these two countries were topping the global statistics together with Italy, Switzerland and Spain with some of the highest infection rates^4^.

**Figure 1A.**
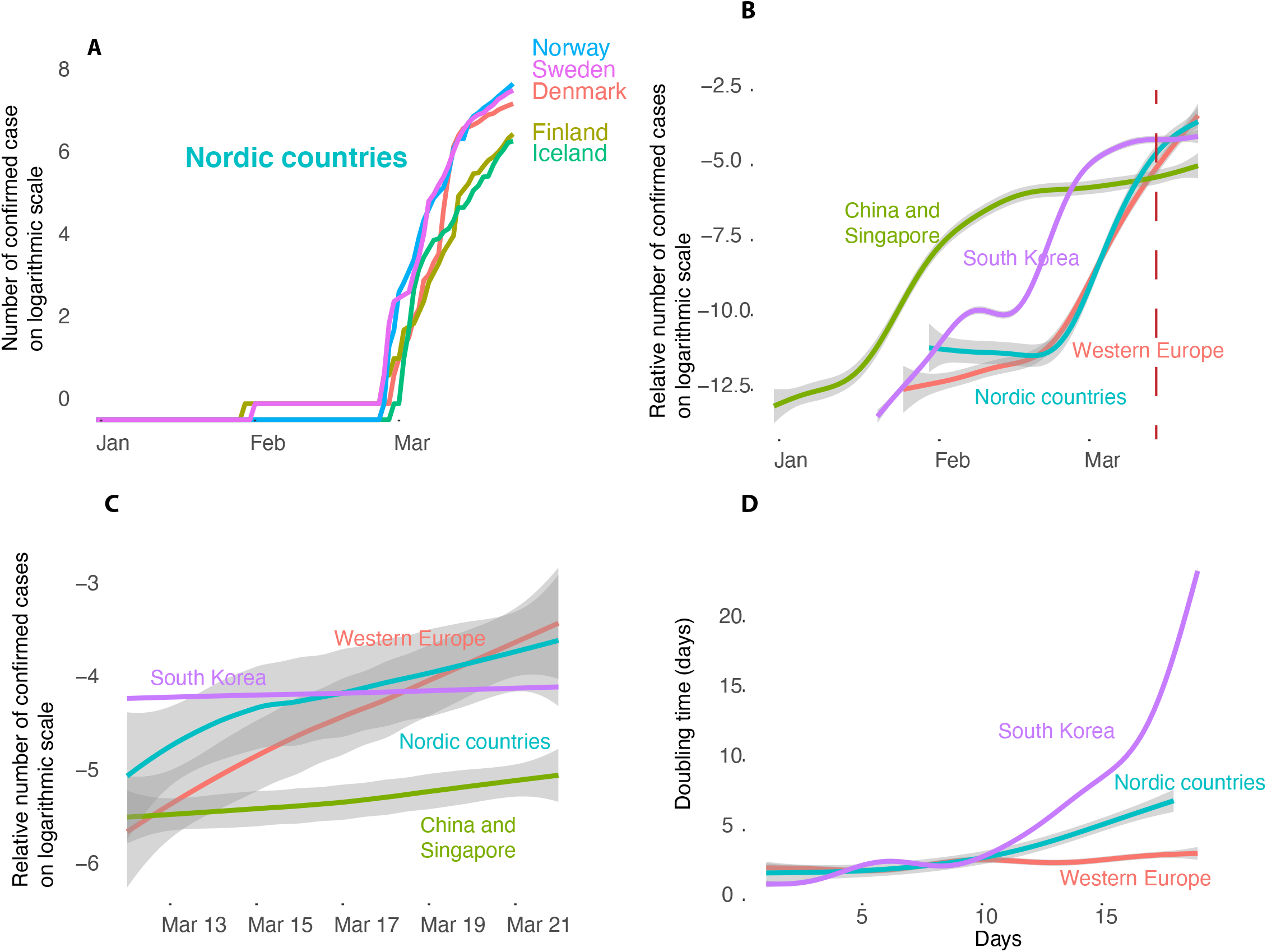
Confirmed COVID-19 cases in Northern Europe Figure 1B. Logarithmic values of the infection rates Figure 1C. Detail from 1B. Figure 1D. Nested DTs plotted day for day. Each DT value was evaluated from a linear model encompassing the start date and the five consequent days. Day 1 in this analysis is approximately the first day of exponential growth. Day 1 in most countries in this study corresponds to Day 60 of the ECDC data set, which was the 29^th^ of February. The exceptions are South Korea (50), Italy (54), Spain (56) and Germany (59). Day “0” in the ECDC data set is the 31st December 2019.

In order to deal with the COVID-19 outbreak, on March 12^th^, 2020 the Norwegian government implemented the most drastic measures since World War 2^5^. Schools and kindergartens were closed, flights were cancelled, public gatherings were postponed and citizens with symptoms were advised to self-quarantine for 14 days. Citizens that had been abroad were required to stay at home with their families for 14 days. On March 16^th^, 2020, the country was officially shut down for travel with strict control of borders. Denmark implemented even stronger measures, with many shops such as IKEA being required to close as well. In the Norwegian capital Oslo, pubs and restaurants are still open, although with a much lower number of visitors than usual. In Norway, companies that could move their employees over to working from home were advised to do so. Iceland and Finland implemented similar measures, while Sweden closed some, but not all, schools. Sweden also recently introduced travel bans.

What has been the impact of shutting down society and encouraging social distancing on the spread of the disease? Are these measures effective?

## Results and discussion

Although the number of confirmed cases is in the range of 2000 for Norway, Sweden and Denmark, and 500 for Iceland and Finland, there is a fear of a lurking “Italian scenario”. Norway has now passed 2400 confirmed cases with 8 deaths (23^rd^ March 2020). The preliminary mortality rates, that indicate the severity of the disease, are currently relatively low for Nordic countries, approximately 1 % or less. According to the Norwegian Institute of Public Health, the majority of confirmed COVID-19 cases were imported by a group of 1000 ski tourists that had returned at the same time from Italy and Austria. This created an “instant explosion” of cases, without being able to confirm a “Patient Zero”.

Statistics following the cumulative number of COVID-19 cases, plotted day for day, do not enable an easy comparison between countries with different population sizes. Moreover, due to steep growths, exponential curves are difficult to compare against each other. We therefore performed comparative analysis on the curves representing infection rates and logarithmic scales for better visualization and modelling (Figure 1A). Figure 1B shows infection rates for Nordic countries when compared to other west European countries. For reference, the statistics for China, Singapore and South Korea were also included.

### Doubling time is changing

From the onset of the epidemic in the Nordic countries to the middle of March, COVID-19 infection rates were similar to those of other West European countries (Figure 1B). This appears to be moderately changing now (Figure 1C). There is an ongoing trend in all Nordic countries that points towards fewer new cases. However, this trend is not easy to follow by looking only at the curves (Figure 1C). One way to follow how the DT trends change is to calculate nested DTs over a period of 5-6 days (Supplemental Figure 1).

Throughout the current epidemic, tremendous efforts have been made in many countries to contain the spread of the virus. The DT values are therefore expected to change over time. A shift towards higher DT values will indicate that the applied measures have been efficient. Figure 1D shows the plot of nested DT values during the exponential growth, comparing Nordic countries with Western Europe and South Korea. The trend in Nordic countries that started shortly after implementing the extraordinary measures for containment of the epidemic on the 12^th^ of March is positive. The increase in DT values is evident in all Nordic countries, especially Denmark. In this country DT changed from 2 to 10 days during few weeks. COVID-19 mortality rates were also relatively low in the Nordic countries so far (Supplemental Figure 2).

### Can we trust the data

A weakness of this and several other studies that are based on publicly available data sets is that it is preliminary and calculated on ongoing data reports during the outbreak of the COVID-19 epidemic. Although the data is curated by the ECDC^6^ and quality is ensured by reporting against the same time of day, small errors in data sets will not be possible to detect. There may be faults in testing COVID-19, as many countries are experiencing difficulties due to a shortage of test kits and a strain on institutions and personnel, which may influence the number of reported cases. South Korea and Norway have both performed a high number of tests. Testing levels in other Scandinavian countries are assumed to be at levels comparable to other West European countries. It is also assumed that the testing routines have been stable for longer periods of time and only change from time to time. To circumvent the testing as a source of error, doubling time of the mortality rates could be calculated as well (Supplemental Figure 3). Cumulative number of deaths plotted on the logarithmic scale shows the same trend as the number of confirmed cases (Supplemental Figure 2).

## Conclusion

Should these results make us feel optimistic about the potential outcomes of COVID-19 in Nordic countries? Realistically, it is still too early to tell. The results support the notion that the decrease of DT in Scandinavia could be the result of governments implementing strict social-distancing measures. The evidence suggests that all Nordic countries which implemented the measures experienced the same trend, which strengthens the hypothesis that the measures are successful. An exception to this is Iceland, where the positive trend towards reduction in the number of new infections started slightly earlier. Another significant factor that may be influencing these outcomes is that certain restrictive measures were already in place in Norway before the 12^th^ of March 2020. People began working from home and travel activities were reduced even before the implementation of official measures.

However positive and encouraging these findings are, it is clear that the DTs in Nordic countries are nowhere close to the corresponding values on the South Korean curve. The exception is Denmark which had DT values of approximately 9-10 days and seems to be fighting the epidemic most strictly. In order to drastically reduce the number of infections in the same way South Korea did, there needs to be more effort on the part of Nordic countries. Should these countries attempt to replicate South Korean example more accurately, the measures currently implemented need to be supplemented by more COVID-19 testing and the widespread use of face masks (which are currently not available).

The experience of Eastern Asian nations indicates that the increase in DT values suggests a favorable scenario with less fatalities. However, there are exceptions; Germany, with average DT below 3 days and 22,000 infected and 84 registered fatalities (23^rd^ March 2020), has one of the lowest mortality rates of 0.3%. In comparison, mortality rates in Italy and UK are 9% and 4.6% respectively. The German exception is likely due to the extensive testing and highly efficient healthcare system in the country. However, there are limitations even to the best systems. A high number of critical patients combined with reduced personnel can overwhelm any country’s healthcare system.

The pleasant Spring weather of the past few days appears to have encouraged many people in Oslo to venture out to city parks, neighboring forests and recreation areas. Collectively, nobody is wearing masks, people are crowding together and are not practicing social distancing. Will this behavior reverse the positive trend that has been building for the past week? From here it can go both ways. Is it going to get worse before it gets better?

The analysis presented here suggests that government-imposed quarantine/shutdown measures are an effective tool for slowing the rate of infection and that these measures need to be sustained and complemented with population-wide testing for maximum impact. Our findings caution against opting for early termination of these policies as is being proposed in the United States.

## Materials and methods

Data analysis was performed in R, open source statistical software. Data set was downloaded from the ECDC site^6^. Linear mathematical models for determination of doubling times were performed in R. Nested DT values for subsequent five days were calculated and plotted. See also (https://medium.com/mindshift/appendix-til-artikkel-i-dagens-medisin-cefe986bb6a) for calculations, data tables and methods.

### UPDATE on March 26^th^, 2020

Parts of this analysis were published in the online magazine, *Today’s Medicine (Dagens Medicine)*, a Norwegian newspaper and a specialist magazine for health professionals, on March 21st^7^. Danish^8^ and Norwegian^9^ Health authorities confirmed observing similar results as well on the 23^rd^ and the 24^th^ March respectively. Updated of this analysis was published on the of 25^th^ of March^10^.

Biljana Stangeland, PhD,

Principal Data Scientist and Biomedical Expert at Mindshift AS, Oslo, Norway

## Data Availability

THe manuscript is based on Open data.

https://www.ecdc.europa.eu/en/geographical-distribution-2019-ncov-cases

## Supplemental Figures

Supplemental Figure 1 Method for calculating nested TDs from the growth curves plotted on the logarithmic scale.

Supplemental Figure 2 Number of COVID-19 infected and the number of deaths on the logarithmic scale

Supplemental Figure 3 DTs calculated for number of deaths for several countries

## Acknowledgements

The authors would like to thank Suzana Petanceska, PhD and Tamara Doncic for critical reading of the manuscript.

## Conflict of interest

None.

## Notes

### Competing Interest Statement

The authors have declared no competing interest.

### Funding Statement

no external funding was received

## References

1. Nowcasting and forecasting the potential domestic and international spread of the 2019-nCoV outbreak originating in Wuhan, China: a modelling study (2020). The Lancet. JT Wu, K Leung and GM Leung, Published online January 31, 2020 https://doi.org/10.1016/S0140-6736(20)30260-9

2. World Health Organization. Coronavirus disease (COVID-2019) situation report 38 (27 Feb 2020). https://www.who.int/docs/default-source/coronaviruse/situation-reports/20200227-sitrep-38-covid-19.pdf?sfvrsn=2db7a09b_4, (23 March, 2020, date last accessed).

3. World Health Organization. Coronavirus disease (COVID-2019) situation report 42 (02 Mar 2020). https://www.who.int/docs/default-source/coronaviruse/situation-reports/20200302-sitrep-42-covid-19.pdf?sfvrsn=224c1add_2, (23 March, 2020, date last accessed).

4. World Health Organization. Coronavirus disease (COVID-2019) situation report 57 (17 Mar 2020). https://www.who.int/docs/default-source/coronaviruse/situation-reports/20200317-sitrep-57-covid-19.pdf?sfvrsn=a26922f2_4, (23 March, 2020, date last accessed).

5. The Government acts to mitigate effects of the COVID-19 pandemic on the economy, https://www.regjeringen.no/no/aktuelt/the-government-acts-to-mitigate-effects-of-the-covid-19-pandemic-on-the-economy/id2693471/

6. European Centre for Disease Prevention and Control, Situation update worldwide, as of 23rd March 2020, https://www.ecdc.europa.eu/en

7. «Doblingstiden har doblet seg – er det grunn til optimisme?» (The doubling time has doubled - is there reason for optimism?), Dagens Medisin, https://www.dagensmedisin.no/artikler/2020/03/21/doblingstiden-har-doblet-seg--er-det-grunn-til-optimisme/

8. «Drastiske tiltak har effekt, mener danske myndigheter» (Danish authorities claim that the drastic measures have an effect), Dagens Medisin, https://www.dagensmedisin.no/artikler/2020/03/24/danske-tiltak-har-effekt/

9. «Ny analyse fra FHI: – Ser klare tegn til at kurven flater ut» (New analysis from the Norwegian Institute of Health: - Clear indications that the curve is flattening), Dagens Medisin, https://www.dagensmedisin.no/artikler/2020/03/24/ny-analyse-fra-fhi--ser-klare-tegn-til-at-kurven-flater-ut/

10. “Mye tyder på at stengte skoler og hjemmekontor virker», (There are indications that the measures, closed schools and home offices, do work), Dagens Medisin, https://www.dagensmedisin.no/artikler/2020/03/25/-mye-tyder-pa-at-stengte-skoler-og-hjemmekontor-virker/

